# Causal analysis of COVID-19 observational data in German districts reveals effects of mobility, awareness, and temperature

**DOI:** 10.1101/2020.07.15.20154476

**Authors:** Edgar Steiger, Tobias Mußgnug, Lars Eric Kroll

**Author notes:** Email addresses (Edgar Steiger), (Tobias Mußgnug), (Lars Eric Kroll).

## Abstract

Mobility, awareness, and weather are suspected to be causal drivers for new cases of COVID-19 infection. Correcting for possible confounders, we estimated their causal effects on reported case numbers. To this end, we used a directed acyclic graph (DAG) as a graphical representation of the hypothesized causal effects of the aforementioned determinants on new reported cases of COVID-19. Based on this, we computed valid adjustment sets of the possible confounding factors. We collected data for Germany from publicly available sources (e.g. Robert Koch Institute, Germany’s National Meteorological Service, Google) for 401 German districts over the period of 15 February to 8 July 2020, and estimated total causal effects based on our DAG analysis by negative binomial regression. Our analysis revealed favorable causal effects of increasing temperature, increased public mobility for essential shopping (grocery and pharmacy), and awareness measured by COVID-19 burden, all of them reducing the outcome of newly reported COVID-19 cases. Conversely, we saw adverse effects of public mobility in retail and recreational areas, awareness measured by searches for “corona” in Google, and higher rainfall, leading to an increase in new COVID-19 cases. This comprehensive causal analysis of a variety of determinants affecting COVID-19 progression gives strong evidence for the driving forces of mobility, public awareness, and temperature, whose implications need to be taken into account for future decisions regarding pandemic management.

## 1. Introduction

As the COVID-19 pandemic progresses, research on mechanisms behind the transmission of SARS-CoV-2 shows conflicting evidence [62, 9, 24]. While effects of mobility have been extensively discussed, less is known on other factors such as changing awareness in the population [26, 37, 67] or the effects of temperature [4, 12, 40]. A limiting factor in many studies is the lack of a causal approach to assess the causal contributions of various factors [23]. This can lead to distorted estimates of the causal factors with observational data [23, 53, 57].

With COVID-19, we find ourselves in a situation in which information on the causal contribution of various influencing factors in the population is urgently needed to inform politicians and health authorities. On the other hand, trials cannot be carried out for obvious ethical and legal reasons. Therefore, when assessing the effects of determinants of SARS-CoV-2 spread, special attention must be paid to strategies for the selection of confounding factors.

Another problem with assessing the effects of various determinants of SARS-CoV-2 spread is the heterogeneity of the countries and regions examined for example in the Johns Hopkins University (JHU) COVID-19 database [7]. The comparison of time series of case numbers from different countries and observational periods can be strongly distorted by different factors like testing capacities and regional variations.

Our objective is to provide estimates of the causal effects of the main drivers of the pandemic with reduced bias. We conducted a scoping review of the available studies regarding signaling pathways and determinants of the spread of SARS-CoV-2 infections and the reported new COVID-19 cases. Then we integrated the current findings into a directed acyclic graph for the progress of the pandemic at the regional level. Using the resulting model and the do-calculus we found identifiable effects without blocked causal paths whose effects can be analyzed with observational data [47]. We used regional time series data of all German districts (401) from various publicly available sources to analyze these questions on a regional level. Germany is a good choice in this regard, because it has ample data on contributing factors on the regional level and has had high testing and treatment capacities from early on in the pandemic.

## 2. Causal Model

We used a directed acyclic graph (DAG) [53, 57] as a tool to analyze the causal relationships between several exposures and SARS-CoV-2 spread. To get an overview on published associations, a scoping review was conducted from 20th to 22nd of May 2020 within Pubmed and Google scholar. Restrictions were applied to English and German language and the publication date in the last one year. The following search terms were applied to abstracts and title in Pubmed (“COVID-19” OR “COVID19” OR “Corona” OR “Coronavirus” OR “SARS-CoV-2”) and connected separately in each case with the exposure variables (“mobility”, “public awareness”, “awareness”, “google trends”,”ambient temperature”, “temperature”). For “mobility”, we analyzed *n* = 8 studies, *N* = 103 were scanned in Pubmed, together with the first ten pages (100 results) in Google scholar (“awareness”/”public awareness”/”google trends” *n* = 9, *N* = 215; “temperature”/”ambient temperature” *n* = 16, *N* = 235). We integrated these findings where possible into the construction of our DAG, which can be seen in Figure 1.

**Figure 1:**
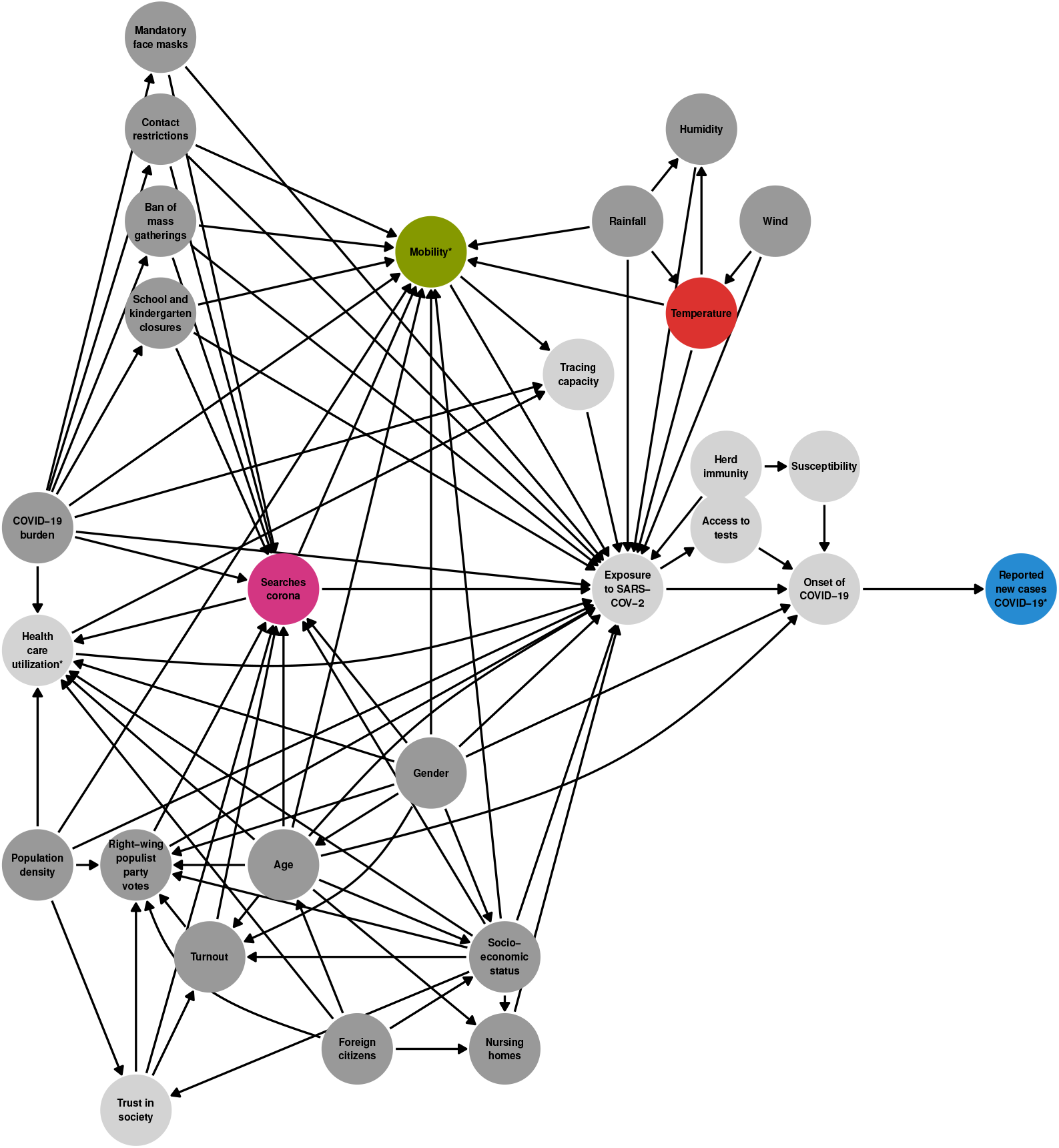
DAG of determinants of reported COVID-19 cases on the district level. Unobserved variables are light gray, variables marked with an asterisk (*) are confounded by weekday/holiday.

A number of studies report a strong association of **mobility** restrictions on the number of new COVID-19 cases: Restrictive measures (e.g. “stay-at-home” orders, travel bans, or school closures) are shown to possibly reduce the COVID-19 incidence [8, 9, 18, 34, 36, 39, 42, 65]. However, some studies point out the combination of various non-pharmaceutical interventions (NPIs) is decisive to prevent new infections [31, 35].

Google Trends [21] data can be used as a tool to get insights into public interest (**awareness**) in the coronavirus disease. Several recent studies imply a connection of relative search volumes (RSV) indices and reported new COVID-19 cases [3, 16, 26, 37, 38, 41, 60, 67, 68]. Some search terms e.g. “COVID-19” or “coronavirus” predated newly infected cases/total number of cases by roughly 7 to 14 days for different countries [16, 26, 37, 67]. Additionally, we acknowledged that individual risk-aware behavior might be a reaction to the current COVID-19 burden (measured as reported cases at the day of exposure).

Mixed evidence is available regarding the effect of **temperature**: On the one hand several papers report an association between increase in temperature and decrease in newly infected COVID-19 cases [4, 12, 40, 50, 54, 55, 58, 61, 63]. On the other hand, also the opposite has been found [2, 64]. Some studies found no association at all [5, 28, 30, 31, 66]. It should be noted that few studies considered other confounding variables than meteorological ones (especially age and population density among others [5, 31, 63]). In addition, the transferability of results between different climate zones is questionable. To avoid possible bias caused by weather variables other than temperature, we included rain, wind, and humidity in our model.

When investigating causal determinants of SARS-CoV-2 infections, a number of confounders have to be considered. Well-known risk factors for SARS-CoV-2 as well as for other infections are demographic factors such as age, gender, socio-economic status (SES), population density, and foreign citizenship/ethnicity [11, 15, 7]. In Germany along with other countries (i.e. Brazil, USA, or the UK), populist parties or politicians and their electorate tend to be more sceptical about effects of containment measures than the other part of the electorate [14, 17]. Therefore we considered both “right-wing populist party votes” and “voter turnout” as possible confounders. Public health interventions were also taken into account (contact restrictions, school closures etc.), as their implementation showed strong correlations with controlling the spread of SARS-CoV-2 [10, 31, 35]. To avoid bias due to reporting delay of case numbers we had to include weekday and German holidays. We include some unobserved variables in our DAG (e.g. “Herd immunity”), too. Please note that “Exposure to SARS-CoV-2” is itself an unobserved variable: German case numbers are reported with delay after date of exposure and symptom onset. *Exposure to the virus* should not be confused with the formal *exposure variables* of the DAG (mobility, awareness, temperature).

## 3. Data

We collected and aggregated data on reported COVID-19 cases, regional socio-demographic factors, weather, and general mobility on district and state level in Germany for the period of 15 February 2020 to 8 July 2020. Our observation period for the outcome consisted of all dates from 23 February 2020 to 8 July 2020 (*T* = 137), since we used a lag of 8 days for all confounders. We did not exclude any states or districts (*K* = 401). We analyzed the daily reported number of new cases as outcome (*K · T* = 54 937 observations). The set of possible predictors was derived from our causal DAG (see Table 1 and Figure 1). Due to modelling and data limitations, some of the predictors were unobserved or were modelled as a construct consisting of several variables. For our causal analysis, we computed adjustment sets in three different scenarios for separate exposures within the DAG: i) mobility of population, ii) awareness of COVID-19 (i.e. Google searches for “corona”), iii) weather (i.e. temperature).

**Table 1:** Observed model variables

| variable | dynamics | level | type | unit/comment | source |
| --- | --- | --- | --- | --- | --- |
| Weekday | daily | national | categorical | Sat through Thu as six binary variables, Fri as baseline | - |
| Holiday (report) | daily | national | binary | - | - |
| Holiday (exposure) | daily | national | binary | - | - |
| **Mobility** |  |  |  |  |  |
| retail and recreation | daily | state | numeric | percent change compared to reference period | Google [20] |
| grocery and pharmacy | daily | state | numeric | percent change compared to reference period | Google [20] |
| parks | daily | state | numeric | percent change compared to reference period | Google [20] |
| workplaces | daily | state | numeric | percent change compared to reference period | Google [20] |
| residential | daily | state | numeric | percent change compared to reference period | Google [20] |
| transit stations | daily | state | numeric | percent change compared to reference period | Google [20] |
| **Weather** |  |  |  |  |  |
| Rainfall | daily | district | numeric | mm (l/sqm) | DWD [13] |
| Temperature | daily | district | numeric | °C | DWD [13] |
| Humidity | daily | district | numeric | relative humidity (%) | DWD [13] |
| Wind | daily | district | numeric | m/s | DWD [13] |
| **Policies** |  |  |  |  |  |
| Ban of mass gatherings | daily | national | binary | - | - |
| School and kindergarten closures | daily | state | numeric | 0 for no closure, 1 for full closure, 0.5 for partial reopening | - |
| Contact restrictions | daily | national | binary | - | - |
| Mandatory face masks | daily | district | binary | - | IZA [43] |
| **Socio-demographic** |  |  |  |  |  |
| Age | constant | district | numeric | 2 variables: share of population $\geq 65$ years & $< 18$ years | INKAR [6] |
| Gender | constant | district | numeric | share of female population | INKAR [6] |
| Population density | constant | district | numeric | population per sqkm | INKAR [6] |
| Foreign citizens | constant | district | numeric | 2 variables: share of foreign citizens & of population seeking refuge | INKAR [6] |
| Socio-economic status | constant | district | numeric | share of households with low income | INKAR [6] |
| Turnout | constant | district | numeric | voter turnout in last election | INKAR [6] |
| Right-wing populist party votes | constant | district | numeric | share of votes for AfD in last election | INKAR [6] |
| Nursing homes | constant | district | numeric | number of nursing (retirement) homes | INKAR [6] |
| **Awareness** |  |  |  |  |  |
| Searches corona | daily | state | numeric | percent relative to other states and observation period | Google [21] |
| COVID-19 burden | daily | district | numeric | reported cases on day of exposure | RKI [52] |
| **Case numbers** |  |  |  |  |  |
| Reported new cases of COVID-19 | daily | district | numeric | - | RKI [52] |
| Active cases | daily | district | numeric | active cases on day of report | RKI [52] |

### 3.1. Variables

We downloaded German daily case numbers on district level reported by Robert Koch Institute (RKI, [52], acquired on 12 July 2020) and aggregated them by date. The number of daily active cases for day *d* was derived by subtracting the total number of reported cases on day *d* and day *d* −14 (14 days as a conservative estimate for the infectious period, which corresponds here to the required quarantine time in Germany).

To assess the mobility of the German population, we used data publicly available on German state level from Google [20]. Measurements are daily relative changes of mobility in percent compared to the period of 3 January 2020 to 6 February 2020. Missing values (25 out of 13 488) were imputed with value 0 and the state level measurements were passed onto districts within the corresponding state. Google mobility data was available for six different sectors of daily life (“retail and recreation”, “grocery and pharmacy”, “parks”, “transit stations”, “workplaces”, “residential”) which means that “mobility” is a construct consisting of several variables. All variables but “residential” mobility are relative changes of daily visitor numbers to the corresponding sectors compared to the reference period. “Residential” mobility is the relative change of daily time spent at residential areas.

The notion of awareness in the population of COVID-19 describes the general state of alertness about the new infectious disease. As such, it was hard to measure directly. As a proxy, we used the relative interest in the topic term “corona” as indicated by Google searches. The daily data was available on state level [21] and passed onto district level. As a second proxy for awareness, we used the daily reported number of COVID-19 cases on the day of the exposure: Since media reported case numbers prominently, we assumed that this could reflect individual awareness, too.

We constructed daily weather from four variables (“temperature”, “rainfall”, “humidity”, “wind”). Weather data was downloaded from Deutscher Wetterdienst (DWD, [13]) for all weather stations in Germany below 1000 meters altitude with daily records for our observation period. District level daily weather data was aggregated per district by averaging the data from the three nearest weather stations (which includes weather stations inside the district). Missing values were imputed with mean values (*n* = 59 for wind).

The reported number of COVID-19 cases varied strongly by day of the week. Thus, we included “weekday” as a categorical variable. Similarly, the reported cases and the exposure to the virus were affected by official holidays. Within the observation period, this included among others Good Friday, Easter Monday, and Labor Day. To correct for effects of these days, we included two variables in the model, “Holiday (report)” (indicates if the day of the report was a holiday, because governmental health departments were less likely to be on full duty) and “Holiday (exposure)” (indicates if the day of exposure to the virus was a holiday, because the population behaves differently on holidays). For different official and political measures we used one-hot encoded daily variables, i.e. ban of mass gatherings, school and kindergarten closures and their gradual reopening, contact restrictions, and mandatory face masks for shopping and public transport.

We included several social, economic, and demographic factors on the district level with direct or indirect influence on the risk of exposure to SARS-CoV-2 in our analysis. All are readily available from INKAR database [6]. We used the share of population that is 65 years or older and the share of population that is younger than 18 years (Age), the share of females in population (Gender), the population density, the share of foreign citizenships and the share of the population seeking refuge (Foreign citizenship), the share of low-income households (Socio-economic status), voter turnout, share of right-wing populist party votes, and the number of nursing (retirement) homes.

All variables but the outcome “Reported new cases of COVID-19” and the offset “Active cases” were centered for numerical stability. We did not scale variables to unit variance to maintain interpretability of effects on the original scale of variables. Additionally, we lagged the effect of all variables (but outcome, offset, and the non-dynamic socio-demographic variables) by 8 days (see Section 5) which means that we assumed that their effects on the outcome will be visible after 8 days.

## 4. Methods

### 4.1. Causal analysis with DAG and adjustment sets

We used a directed acyclic graph as a graphical representation of the hypothesized causal reasoning that leads to exposure to the SARS-CoV-2 virus, onset of COVID-19, and finally reports of COVID-19 cases. Every node *v*_*i*_ in the graph is the graphical representation of an observed or unobserved variable *x*_*i*_, a directed edge *e*_*ij*_ is an arrow from node *v*_*i*_ to *v*_*j*_ that implies a direct causal relationship from variable *x*_*i*_ onto variable *x*_*j*_. The set of all nodes is denoted by *V*, the set of all edges by *E*, as such, the complete DAG is the tuple *G* = (*V, E*). The seminal works of Spirtes and Pearl [56, 46] introduce the theory of causal analysis, do-calculus, and how to analyze a DAG to estimate the total or direct causal effect from a variable *x*_*i*_ onto a variable *x*_*j*_. The direct effect is the effect associated with the edge *e*_*ij*_ only (if it exists), while the total effect takes indirect effects via other paths from *v*_*i*_ to *v*_*j*_ into account, too. Here we estimated total effects only, since most of our variables were not hypothesized to have a direct effect on the *reported* number of new COVID-19 cases. In contrast to prediction tasks, where one would include all variables available, it is actually ill-advised to use all available variables to estimate causal effects, due to introducing bias by adjusting for unnecessary variables within the causal DAG. This is why we need to identify a valid set of necessary variables (an adjustment set) to estimate the proper causal effect [46]. The “minimal adjustment set” [22] is a valid adjustment set of variables that does not contain another valid adjustment set as a subset. However, identifying a minimal adjustment set might not be enough to reliably estimate the causal effect. Thus, we identified the “optimal adjustment set” [25] as the set of variables which is a valid adjustment set while having the lowest asymptotic variance in the resulting causal effect estimates.

We analyzed the DAG from Section 2 with the R Software [51] and the R packages dagitty (formal representation of the graph and minimal adjustment sets [57]) and pcalg (for finding an optimal adjustment set [32]). For the defined exposures and the outcome “Reported new cases of COVID-19”, we computed the minimal and optimal adjustment sets. Since it was possible that these sets contained unobserved variables that needed to be left out of the regression model, we chose the valid set with the highest pseudo-*R*^2^ (see next section) to estimate the final total causal effect from exposure to outcome.

### 4.2. Regression with negative binomial model

We can estimate the causal effect from exposure to outcome by regression [46]. Since the outcome “Reported new cases of COVID-19” is a count variable, one should not employ a linear regression model with Gaussian errors, but instead we assumed a log-linear relationship between the expected value of the outcome *Y* (new cases) and regressors *x*, as well as a Poisson or negative binomial distribution for *Y* :

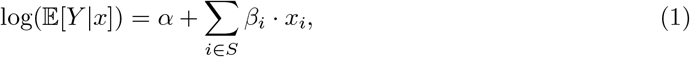

where *α* is the regression intercept, *S* is the set of adjustment variables for the exposure *i*^*\**^ including the exposure variable itself, *β*_*i*_ are the regression coefficients corresponding to the variables *x*_*i*_. As such 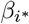 is the total causal effect from exposure variable 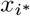 on the outcome Y.

The Poisson regression assumes equality of mean and variance. If this is not the case one observes so-called overdispersion (the variance is higher than the mean), this indicates one should use regression with a negative binomial distribution instead to estimate the variance parameter separately from the mean.

We needed to account for the fact that our outcome is not counted per time unit (one day) only, but depends on the number of active COVID-19 cases: Holding all other variables fixed, the number of new cases *Y* is a constant proportion of the number of active cases *A*. This was modeled by including an offset log(*A* + 1) in the regression model (1):

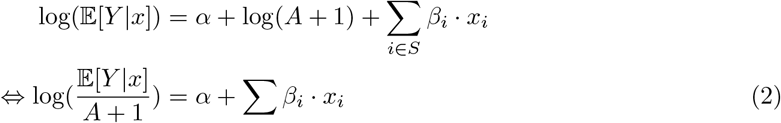

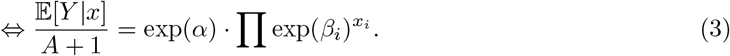

Here we added a pseudocount “+1” to ensure a finite logarithm and avoid division by 0.

One can interpret the model as approximating the log-ratio of new cases and active cases by a linear combination of the regressor variables (2). If all variables *x*_*i*_ are centered in (3), we have for the baseline *∀i x*_*i*_ = 0 *⇒*𝔼 [*Y* |*x* = 0] = exp(*α*) · (*A* + 1). In other words, the exponentiated intercept is the baseline daily infection rate (how many people does one infected individual infect in one day). If we hold all variables *x*_*i*_ fixed (e.g. at baseline 0) in (3) but now increase the exposure variable *x*_*i\**_ = 0 by one unit to 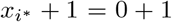, we have 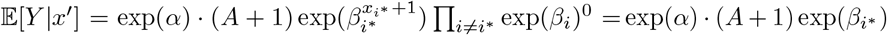, which means the exponentiated coefficient 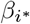 describes the rate change of the outcome by one unit increase of the exposure.

In practice, given observations of *Y* and *x* we estimate the regression coefficients *α* and *β*_*i*_ by *maximum likelihood* [27]. Our observational measurements are *y*_*kt*_ and *x*_*ikt*_, where *k* indicates the corresponding district and *t* the date of measurement.

When we analyzed different adjustment sets given by analysis of the causal DAG (i.e. the minimal and optimal adjustment sets), we first checked if the set included unobserved variables. If this was the case for the optimal adjustment set, we discarded the unobserved variables from the set and checked if it was still a valid adjustment set (function gac in package pcalg [48]). If a minimal adjustment set contained unobserved variables, we discarded the whole set. We conducted a log-linear regression (function glm with family=poisson() for Poisson regression, and glm.nb from the MASS package for the negative binomial regression [59]) for every remaining valid adjustment set as regressors and calculated a Pseudo-*R*^2^ given by 1 − *V*_*m*_*/V*_0_, where *V*_*m*_ is the sum of squared prediction errors of the current model and *V*_0_ is the sum of squared prediction errors of the null model (intercept and offset only). That is, our Pseudo-*R*^2^ is 1 minus the fraction of variance unexplained. Finally, we decided for the model/adjustment set with the highest pseudo-*R*^2^. We report the exponentiated estimated coefficients along with 99 percent confidence intervals of the estimates.

## 5. Results

Descriptive statistics for the included variables are presented in Table 2.

**Table 2:**
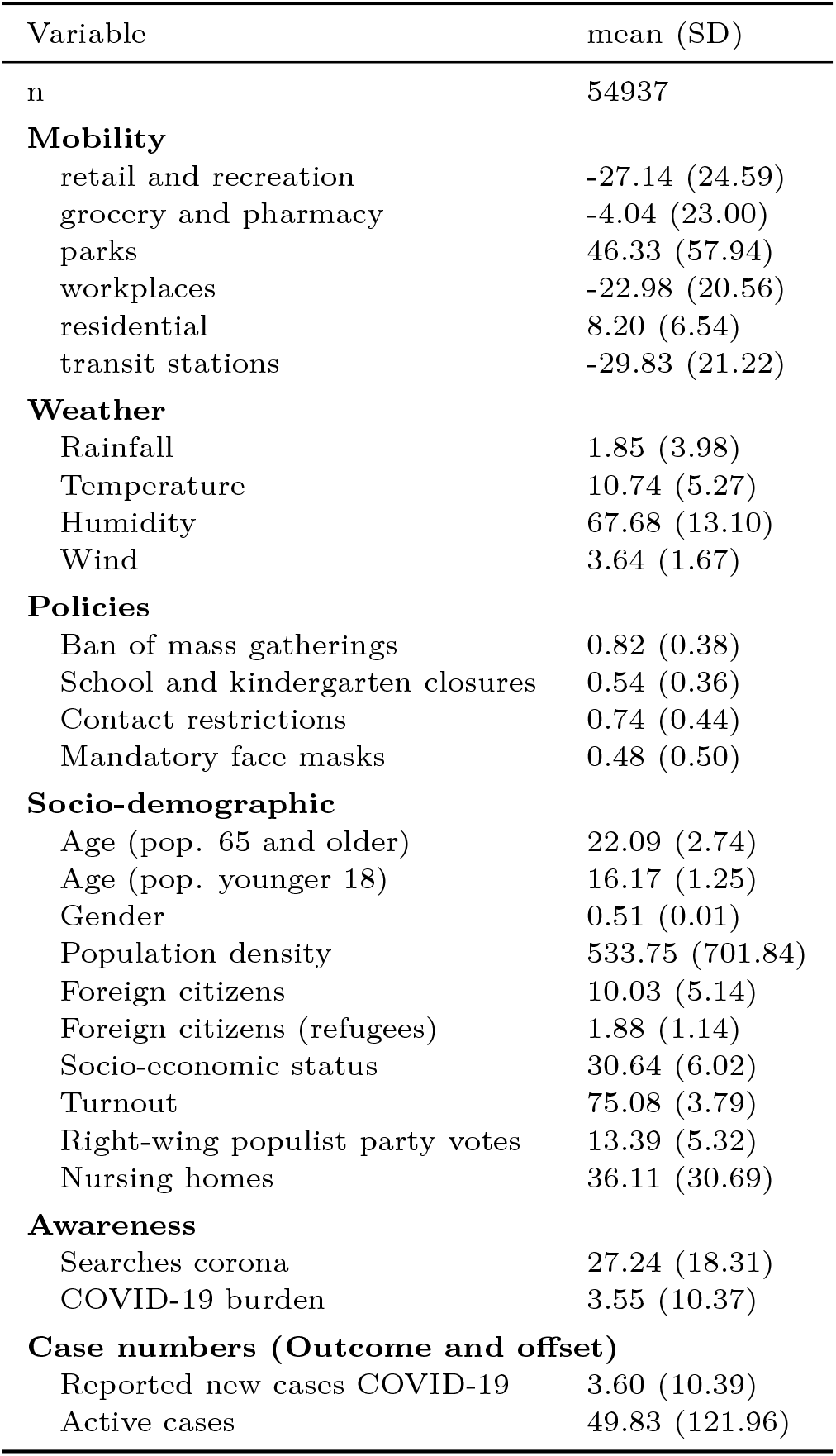
Descriptive Statistics for observed variables

In the observational period, the number of daily reported COVID-19 cases increased till the end of March/beginning of April and continually decreased afterwards till the beginning of June 2020 with a slight increase and decrease afterwards (Figure 2A). On the other hand, the (log-)ratio of reported cases over active cases decreased steeply till the mid of April and increased steadily afterwards with a slight decrease close to the end of the observation period (Figure 2B). Both figures examplify a considerable variation among the districts (light blue points are individual district’s data).

**Figure 2:**
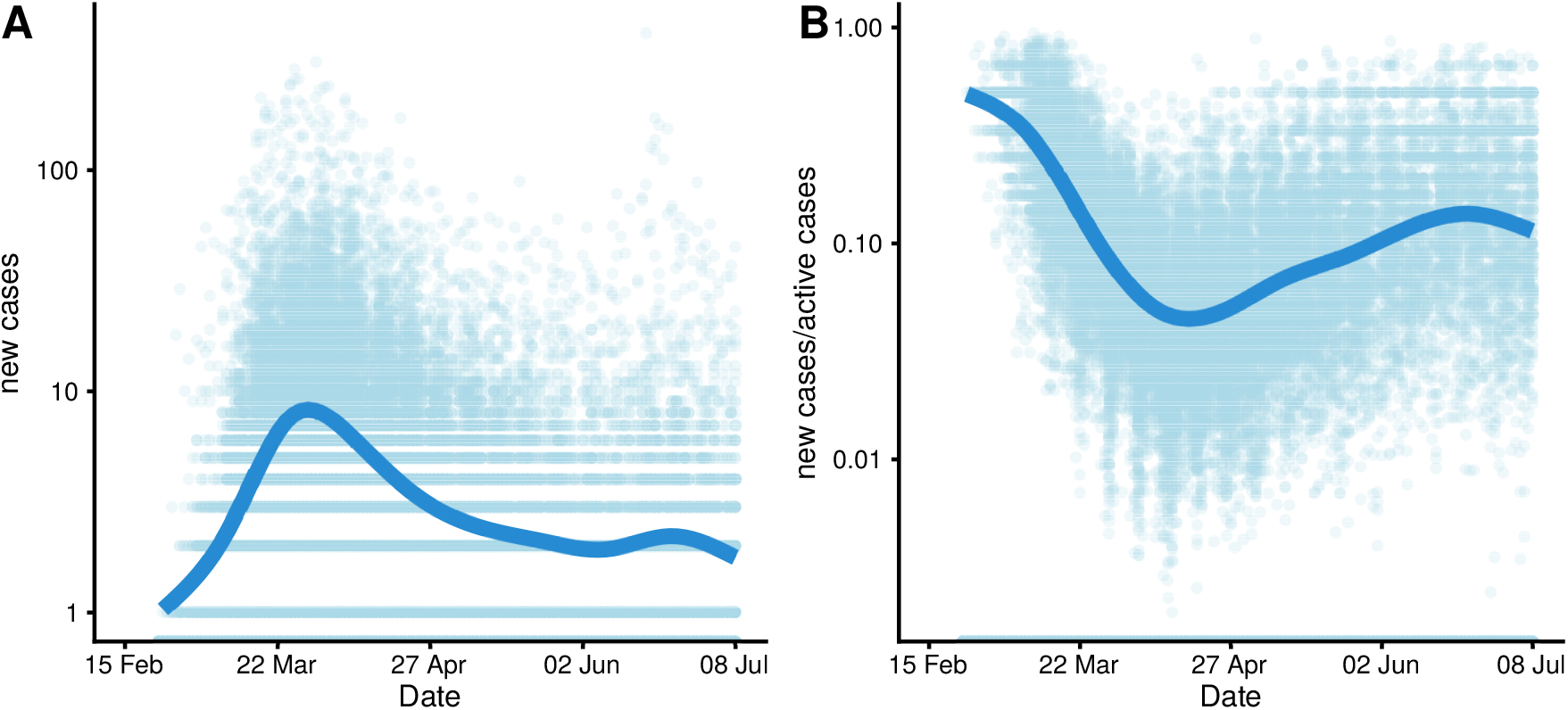
Temporal and district level variation of outcome (log-scale)

In Germany, we observed a rebound in mobility after the initial political measures, reductions in incident cases were associated with a diminishing public interest in COVID-19, and temperatures were overall increasing (cf. Figure 3); with correlations between temporal progression and mobility in retail and recreation *r*_*A,D*_ = −0.02, awareness (“Searches corona”) *r*_*A,C*_ = −0.29, and temperature *r*_*A,B*_ = 0.79.

**Figure 3:**
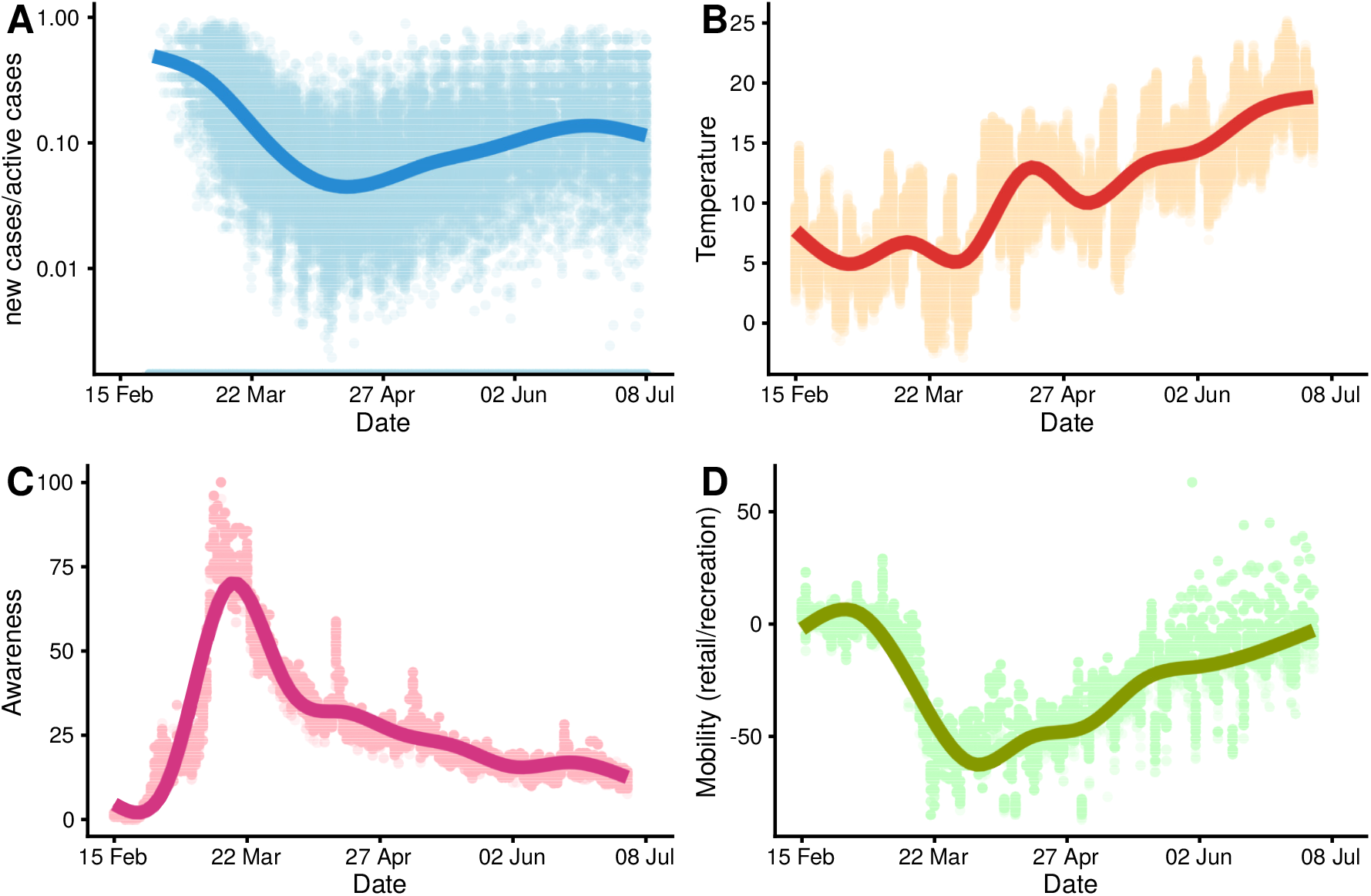
Temporal variation of outcome and main determinants

### 5.1. Main results

We list the results of our causal analysis for the effects of our variables in Table 3. The estimates are multiplicative rates of increase/decrease for a one unit increase of the respective variable: Values above 1 lead to an increase, below 1 to a decrease of the infection rate. To put these estimates into perspective, Figure 4 shows the relative causal effect of the different exposure variables on the number of reported COVID-19 cases on a range of sensible values of the exposure variables (95 percent quantiles of data points).

**Table 3:** Causal effect estimates with 99 percent confidence bands

| cause | effect estimate | CI (lower) | CI (upper) |
| --- | --- | --- | --- |
| **Mobility** |  |  |  |
| retail and recreation | 1.0076 | 1.0056 | 1.0097 |
| grocery and pharmacy | 0.9951 | 0.9938 | 0.9965 |
| parks | 0.9998 | 0.9994 | 1.0003 |
| transit stations | 0.9975 | 0.9946 | 1.0004 |
| workplaces | 1.0031 | 1.0002 | 1.0059 |
| residential | 1.0080 | 0.9987 | 1.0174 |
| **Awareness** |  |  |  |
| Searches corona | 1.0101 | 1.0085 | 1.0118 |
| COVID-19 burden | 0.9942 | 0.9932 | 0.9951 |
| **Weather** |  |  |  |
| Temperature | 0.9918 | 0.9879 | 0.9958 |
| Rainfall | 1.0108 | 1.0066 | 1.0150 |
| Humidity | 1.0022 | 1.0006 | 1.0039 |
| Wind | 0.9893 | 0.9802 | 0.9985 |

**Figure 4:**
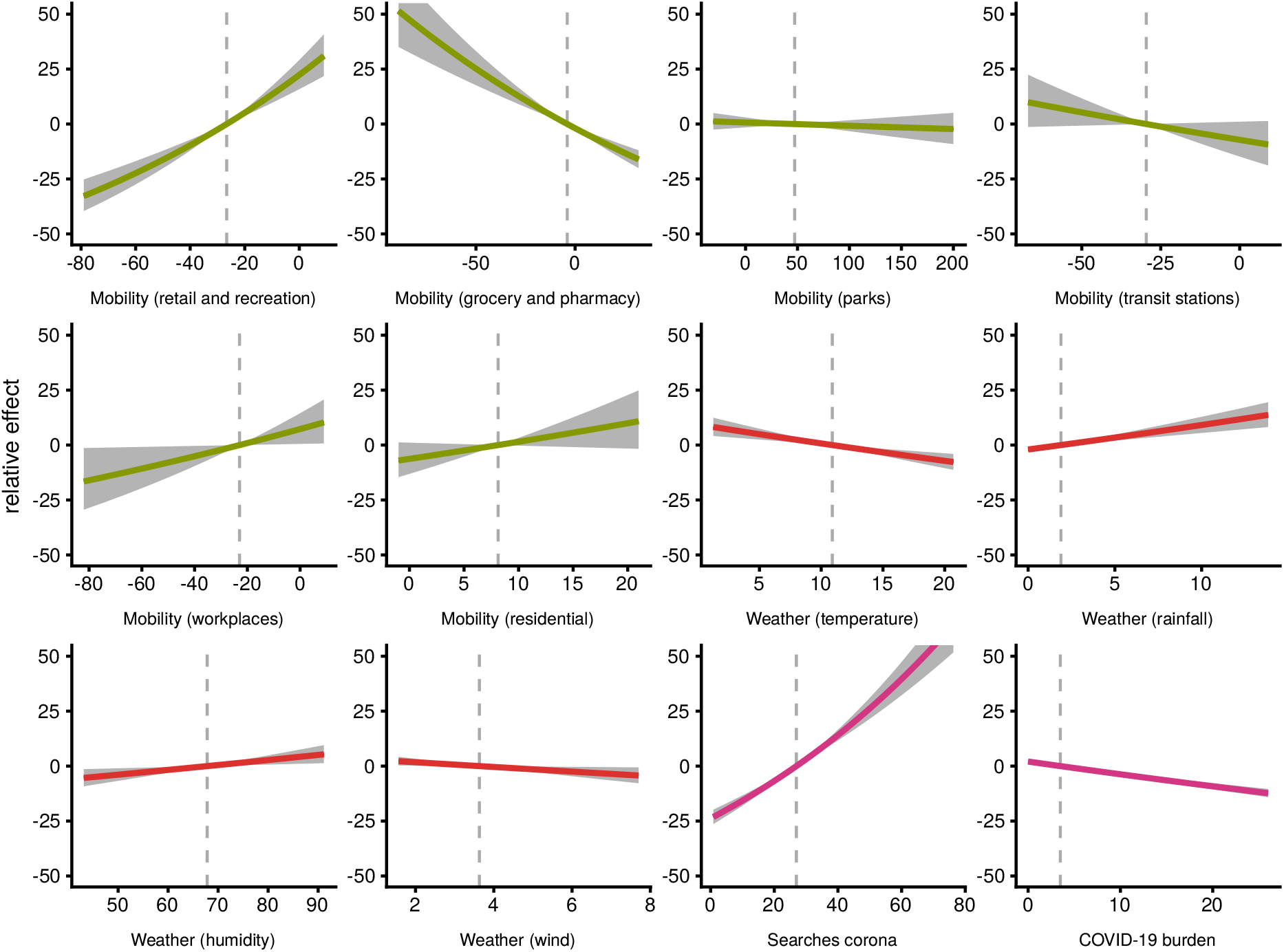
Relative causal effects of exposures

Within our framework, we saw significant effects for mobility in retail/recreational areas and essential shopping (grocery and pharmacy). For retail/recreation, an increase of 1 percent point mobility compared to the reference period (03 January to 06 February 2020) leads to an increase of the daily reported case number by about 0.8 percent. Contrarily, a corresponding increase of 1 percent point for the areas of grocery/pharmacy leads to a decrease in the reported case number by approximately 0.5 percent. Mobility on workplaces showed a small effect of 0.3 increase in case numbers for every 1 percent point increase in mobility. Other causal effects of mobility were insubstantial and not consistent in their direction (99 percent confidence intervals of estimates include 1). Figure 4 shows the effects of mobility on a range of possible values. Thus, we expect an increase of daily cases by approximately 23 percent if mobility in retail/recreation reaches baseline levels of 0 percent difference to the reference period. On the other hand, an increase of mobility for grocery/pharmacy by 10 percent points compared to the reference period leads to a reduction of the infection rate by approximately 7 percent.

“Awareness” had two opposite effects on the outcome in our DAG. Awareness measured by Google searches for *corona* had a positive effect on the number of reported cases. An one percent point increase of the state’s Google searches (relative to other states and the observation period) leads to an increase of approximately 1 percent. For example, if a district shows 10 percent points more relative searches for *corona* than another one, we expect approximately 11 percent more infections for this district after 8 days. *COVID-19 burden* (reported number of cases on day of exposure) affected the outcome negatively, where every additional daily case in the district leads to a 0.6 percent decrease in newly reported case numbers. The last plot in Figure 4 visualizes this relationship: For a local outbreak with 25 daily cases as COVID-19 burden, we estimate as total causal effect a subsequent reduction of infection rate by 11.8 percent.

Within our model, we observed a causal effect of temperature and all other weather variables. Every increase of 1 degree Celsius in temperature leads to a reduction of the daily reported case numbers by approximately 0.8 percent. On the other hand, we found an increasing effect of rainfall: One millimeter (=1 liter per square meter) more rainfall leads to an increase of reported case numbers by approximately 1.1 percent. We observe effects for humidity and wind as well (higher humidity leading to more cases, stronger wind leading to less cases). In perspective (Figure 4), with temperature we expect an increase by approximately 9.2 percent at a daily average temperature of 0*°C*. For rainfall, we expect on a rainy day with 10 mm rainfall a corresponding increase of the infection rate by approximately 9.2 percent.

In all cases we opted to use the reduced optimal adjustment set over the minimal adjustment sets because of higher pseudo-*R*^2^ values (mostly above 0.3), except for mobility, where the minimal adjustment set had a higher pseudo-*R*^2^. Notably, these sets always include most of our socio-demographic variables as confounders as well as the policy variables (cf. Table 4, with the exception being COVID-19 burden). We also decided for the lag of 8 days based on the highest pseudo-*R*^2^ values compared to other lags on the chosen adjustment sets. Similarly, negative binomial regression was chosen over Poisson regression, because the latter showed overdispersion and overall lower pseudo-*R*^2^ values.

**Table 4:** Final adjustment sets for causal analysis

|  | Mobility | COVID-19 burden | Searches corona | Temperature | Rainfall | Humidity | Wind |
| --- | --- | --- | --- | --- | --- | --- | --- |
| Pseudo-R2 | 0.332 | 0.151 | 0.298 | 0.298 | 0.286 | 0.306 | 0.286 |
| Weekday | x | x | x | x | x | x | x |
| Holiday (report) | x | x | x | x | x | x | x |
| Holiday (exposure) | x | x | x | x | x | x | x |
| **Mobility** |  |  |  |  |  |  |  |
| retail and recreation |  |  |  |  |  |  |  |
| grocery and pharmacy |  |  |  |  |  |  |  |
| parks |  |  |  |  |  |  |  |
| workplaces |  |  |  |  |  |  |  |
| residential |  |  |  |  |  |  |  |
| transit stations |  |  |  |  |  |  |  |
| **Weather** |  |  |  |  |  |  |  |
| Rainfall | x | x | x | x |  | x | x |
| Temperature | x | x | x |  |  | x |  |
| Humidity |  | x | x |  |  |  |  |
| Wind |  | x | x | x | x | x |  |
| **Policies** |  |  |  |  |  |  |  |
| Ban of mass gatherings | x |  | x | x | x | x | x |
| School and kindergarten closures | x |  | x | x | x | x | x |
| Contact restrictions | x |  | x | x | x | x | x |
| Mandatory face masks |  |  | x | x | x | x | x |
| **Socio-demographic** |  |  |  |  |  |  |  |
| Age | x | x | x | x | x | x | x |
| Gender | x | x | x | x | x | x | x |
| Population density | x | x | x | x | x | x | x |
| Foreign citizens |  | x | x |  |  |  |  |
| Socio-economic status | x | x | x | x | x | x | x |
| Turnout |  | x |  |  |  |  |  |
| Right-wing populist party votes |  | x | x | x | x | x | x |
| Nursing homes |  | x | x | x | x | x | x |
| **Awareness** |  |  |  |  |  |  |  |
| Searches corona | x |  |  | x | x | x | x |
| COVID-19 burden | x |  | x | x | x | x | x |

## 6. Discussion

### 6.1. Main findings

Our objective was to identify causal effects for COVID-19 cases. We found that weather affects the reported number of infections, especially temperature (which has a reducing effect on case numbers) and rainfall (which increases case numbers). We saw that reports of high case numbers in districts led to a reduction in new infection numbers, which indicates risk-averse awareness in the population and/or effective public health measures to suppress a local outbreak. The overall effect of mobility showed no consistent effect, however, in specific areas significant causal effects could be measured: Increasing activity in retail and recreational areas increased reported case numbers, while increased movement for essential shopping (grocery and pharmacy) led to reduced case numbers.

Furthermore, we made a strong case for the use of causal DAGs in epidemiology and a pandemic like COVID-19: DAGs allow to choose confounders for the analysis in a principled and statistically correct way while reducing possible causes for bias. Also, the DAG formalization allows for discussion about the underlying causal assumptions.

### 6.2. Comparison with previous research

Most research on determinants affecting case numbers of COVID-19 is restricted to single aspects [18, 37, 54, 61]. To reliably identify causal drivers, one must adjust for confounders. To this end, we used an integrated model with variables from different aspects like mobility, awareness, weather, or socio-demographics and identified confounders by causal analysis with a directed acyclic graph. A causal approach is used in another current COVID-19 analysis [19]. There, however, they identify the causal relationships (reconstruct a DAG), while we estimated causal effects for a given hypothesized DAG.

Several studies assessing the impact of public health measures on mobility have each observed a downward trend accompanied by a decrease in the number of newly reported cases [8, 10, 18, 34, 35, 39].

Our findings regarding awareness/Google Trends analysis are in good agreement with the correlations found by Effenberger et al. [16], Higgins et al. [26], and Yuan et al. [67], who conclude that alertness to COVID-19 rises several days before the highest number of cases are reported. At this point it should be noted, that awareness is substantially influenced by public media coverage, which should be considered, if possible, in future studies [26]. As such, awareness is difficult to measure and here the number of Google searches for “corona” could only be a proxy for this concept. In addition, in alignment with other recent published studies, our results confirm evidence which associated a negative effect of temperature on new COVID-19 cases [4, 12, 40, 50, 54, 55, 58, 61, 63]. It is however controversial to other scientific literature describing no effects [5, 28, 30, 31, 66] or even converse correlations [2, 64]. The conflicting results might be explained by different climates and characteristics of the populations under study. While we are confident that our strict causal analysis resulted in effect estimates as undistorted as possible, there might be unconsidered bias in those other studies. Further research needs to be done to elucidate the biological characteristics of the novel virus SARS-CoV-2 regarding its ambient temperature survival and transmission. Finally, we found a positive causal effect of increment precipitation and a raise in COVID-19 cases, which supports previous observations [55].

### 6.3. Limitations and strengths

While use of a causal DAG is itself a strong tool to identify causal effects (and not just statistical associations), it introduces two limitations: causal assumptions within the graph (depicted by edges) need to be well justified, and the statistical regression model that calculates total causal effects needs to be appropriate for the task at hand. We endorse our graph as a basis for discussion on residual confounding. We did not try to construct the DAG from the available data (cf. [19]). As such, our proposed DAG is not entirely consistent with the data and there are conditional dependencies between variables that cannot be dissolved by adding edges to the DAG (e.g. between the policies like contact restrictions and mandatory face masks). Another way to identify potential problems in the proposed DAG is to perform a sensitivity analysis of its structure by inspecting its maximal ancestral graph (MAG) or its Markov equivalence class represented by a complete partially DAG (CPDAG) and the existence of valid adjustment sets for these generalized graphs [49]. For the MAG derived from our DAG, only the effects for exposures mobility and searches for corona can be estimated with valid adjustment sets, while for the Markov equivalence class all exposures but COVID-19 burden lead to valid adjustments sets. A further analysis of these implications is out of the scope of this paper.

We observed overdispersion and a substantial increase in model performance with a negative binomial regression compared to Poisson regression, which is in line with the results on COVID-19 daily case counts of Kraemer et al. [34] and others [40, 4, 29]. We did not model case counts with a differential equation model like the classic SIR-model [33] and its successors, since these are more suited to prediction [e.g. 1] while our choice of a negative binomial regression framework allowed us to estimate the effects of confounders more reliably. There are more advanced statistical methods for count data, e.g. zero-inflated models and mixed models. We tested both approaches as extensions to the negative binomial regression and experienced numerical problems and increased computing time, along with an insubstantial increase in model performance. Furthermore, our model assumed that all variables have effects proportional to the size of their measurements. It is possible that some variables show saturation effects or opposite effects for low, medium, or high values. This could be modelled with polynomial or other transformations of the variables, which we did not employ due to limited temporal and spatial data availability. Use of a fixed DAG with effect estimation via regression assumes that data was generated by the same underlying process for the observation period. By inclusion of the successive mitigation policies as binary variables we were able to explain some of the variance caused by the changing dynamics of case numbers (similar to [29]).

We stress the point that our effects were deduced on an aggregate (district) level in the absence of available data on an individual level. As such, conclusions about effects cannot be transferred on individuals without the possibility for an ecological fallacy. Furthermore, as we were using administrative data for our analysis, the results are susceptible to the Modifiable Area Unit Problem (MAUP) [44]. The MAUP postulates that different regional aggregations of the units of observation may lead to different results and conclusions. Due to limited available data for the different variables, there is currently no way to overcome these problems that are inherent to all analyses on aggregated data level.

Our observation period was restricted to succession from late winter to spring and summer (February to July). Nevertheless, this transition with increasing temperature was a natural experiment that allowed clues on weather effects.

We could not include data on health care utilization during the pandemic into our models due to the lack of available resources. This is planned for a later follow up to this paper since we rank health care utilization and mobility within health care facilities among the strong factors for COVID-19 progression: personnel in hospitals and private practices is particularly exposed to infection, while the lack of adequate care for other diseases has severe effects on general health of the population. At the same time, health care facilities are key for testing and surveillance of COVID-19 patients.

While our analysis focused on Germany and its districts, we assume that results may be transferred to other countries by adjusting for their respective weather conditions, mobility habits, socio-demographic characteristics, and other determining factors.

The code and resources for our analysis are available on Github, we invite other researchers to replicate our analysis with different assumptions using the files provided in the repository^1^ of the article.

### 6.4. Discussion of causal effects

In our analysis, the adverse effect of mobility in retail/recreation and the favorable effect of mobility in grocery/pharmacy indicate that policies like contact restrictions which limit the number of individual interactions can lead to reduced infection numbers. This is due to retail/recreational areas encompassing mostly places of social gatherings like restaurants and bars, malls, sports and music venues, among others, while if people are doing more of their essential shopping at supermarkets, they will most likely stay at home with less contact to other people.

The causal effects of awareness measured via searches for “corona” and the COVID-19 burden are harder to interpret. We assume that within our model, the searches for “corona” are an insufficient proxy for awareness, while the decreasing effect for future case numbers of high daily COVID-19 burden indicates it affects individual risk-behavior and entails effective non-pharmaceutical interventions.

Similarly, the effects of temperature and rainfall can be interpreted as causal effects for indoor and outdoor activities, such that higher temperatures and low rainfall indicate more people spending time outdoor while lower temperatures and high rainfall result in indoor activities, which lead to more infections. Current research suggests this to be due to the prevalent airborne and respiratory droplets and aerosol transmission of the SARS-CoV-2 virus [45]. In this light, we advocate for precautious measures like increased hygiene, face masks, and air ventilation for unavoidable indoor activities.

### 6.5. Conclusions

To the best of our knowledge, this is the most comprehensive analysis of causes for COVID-19 infections which integrates different data sources (all publicly available). Causal reasoning with a DAG allows us to estimate the causal effects more reliably.

Our findings suggest that the causal effects of mobility, awareness, and weather need to be taken strongly into account when deciding for mitigation and suppression measures, depending on the recent and future COVID-19 pandemic development.

## Data Availability

All data gathered from publicly available sources, see Github repository pertaining to the manuscript.

https://github.com/zidatalab/causalcovid19

## Acknowledgments

We are thankful for feedback from Thomas Czihal, Johannes Textor, and Ralph Brinks, who gave helpful suggestions on an earlier draft of the manuscript.

## Footnotes

1 The repository is located here: https://github.com/zidatalab/causalcovid19

## Notes

### Competing Interest Statement

The authors have declared no competing interest.

### Funding Statement

No external funding.

### Author Declarations

not applicable (observational study on publicly available data)

## References

[1] Matthias an der Heiden and Udo Buchholz. Modellierung von Beispielszenarien der SARS-CoV-2-Epidemie 2020 in Deutschland. 2020. doi:10.25646/6571.2.

[2] A.C.F. Auler, A. M. Cássaro, V. O. da Silva, and L. F. Pires. Evidence that high temperatures and intermediate relative humidity might favor the spread of COVID-19 in tropical climate: A case study for the most affected Brazilian cities. The Science of the total environment, 729: 139090, April 2020. ISSN 1879-1026 0048-9697 0048-9697. doi:10.1016/j.scitotenv.2020.139090.

[3] Seyed Mohammad Ayyoubzadeh, Seyed Mehdi Ayyoubzadeh, Hoda Zahedi, Mahnaz Ahmadi, and Sharareh R Niakan Kalhori. Predicting COVID-19 Incidence Through Analysis of Google Trends Data in Iran: Data Mining and Deep Learning Pilot Study. JMIR Public Health and Surveillance, 6(2):e18828, April 2020. ISSN 2369-2960 2369-2960. doi:10.2196/18828.

[4] Melanie Bannister-Tyrrell, Anne Meyer, Celine Faverjon, and Angus Cameron. Preliminary evidence that higher temperatures are associated with lower incidence of COVID-19, for cases reported globally up to 29th February 2020. medRxiv, page 2020.03.18.20036731, January 2020. doi:10.1101/2020.03.18.20036731. URL http://medrxiv.org/content/early/2020/03/20/2020.03.18.20036731.abstract.

[5] Álvaro Briz-Redón and Ángel Serrano-Aroca. A spatio-temporal analysis for exploring the effect of temperature on COVID-19 early evolution in Spain. The Science of the total environment, 728: 138811, April 2020. ISSN 1879-1026 0048-9697 0048-9697. doi:10.1016/j.scitotenv.2020.138811.

[6] Bundesinstitut für Bau-, Stadt- und Raumforschung (BBSR). INKAR – Indikatoren und Karten zur Raumund Stadtentwicklung, 2020, accessed 2020-06-25. URL https://www.inkar.de/.

[7] Center for Systems Science and Engineering (CSSE). COVID-19 Data Repository by the Center for Systems Science and Engineering (CSSE) at Johns Hopkins University, 2020. URL https://github.com/CSSEGISandData/COVID-19.

[8] Meng-Chun Chang, Rebecca Kahn, Yu-An Li, Cheng-Sheng Lee, Caroline O Buckee, and Hsiao-Han Chang. Modeling the impact of human mobility and travel restrictions on the potential spread of SARS-CoV-2 in Taiwan. medRxiv, page 2020.04.07.20053439, January 2020. doi: 10.1101/2020.04.07.20053439. URL http://medrxiv.org/content/early/2020/04/11/2020.04.07.20053439.abstract.

[9] Matteo Chinazzi, Jessica T. Davis, Marco Ajelli, Corrado Gioannini, Maria Litvinova, Stefano Merler, Ana Pastore y Piontti, Kunpeng Mu, Luca Rossi, Kaiyuan Sun, Cécile Viboud, Xinyue Xiong, Hongjie Yu, M. Elizabeth Halloran, Ira M. Longini, and Alessandro Vespignani. The effect of travel restrictions on the spread of the 2019 novel coronavirus (COVID-19) outbreak. Science, 368(6489):395–400, 2020. ISSN 0036-8075. doi: 10.1126/science.aba9757. URL https://science.sciencemag.org/content/368/6489/395.

[10] Benjamin J Cowling, Sheikh Taslim Ali, Tiffany W Y Ng, Tim K Tsang, Julian C M Li, Min Whui Fong, Qiuyan Liao, Mike YW Kwan, So Lun Lee, Susan S Chiu, Joseph T Wu, Peng Wu, and Gabriel M Leung. Impact assessment of non-pharmaceutical interventions against coronavirus disease 2019 and influenza in Hong Kong: an observational study. The Lancet Public Health, 5(5):e279–e288, May 2020. ISSN 2468-2667. doi: 10.1016/S2468-2667(20)30090-6. URL https://doi.org/10.1016/S2468-2667(20)30090-6.

[11] Simon de Lusignan, Jienchi Dorward, Ana Correa, Nicholas Jones, Oluwafunmi Akinyemi, Gayatri Amirthalingam, Nick Andrews, Rachel Byford, Gavin Dabrera, Alex Elliot, Joanna Ellis, Filipa Ferreira, Jamie Lopez Bernal, Cecilia Okusi, Mary Ramsay, Julian Sherlock, Gillian Smith, John Williams, Gary Howsam, Maria Zambon, Mark Joy, and F D Richard Hobbs. Risk factors for SARS-CoV-2 among patients in the Oxford Royal College of General Practitioners Research and Surveillance Centre primary care network: a cross-sectional study. The Lancet Infectious Diseases. ISSN 1473-3099. doi: 10.1016/S1473-3099(20)30371-6. URL https://doi.org/10.1016/S1473-3099(20)30371-6.

[12] Jacques Demongeot, Yannis Flet-Berliac, and Hervé Seligmann. Temperature Decreases Spread Parameters of the New Covid-19 Case Dynamics. Biology, 9(5), May 2020. ISSN 2079-7737 2079-7737. doi: 10.3390/biology9050094.

[13] Deutscher Wetterdienst (DWD) Climate Data Center (CDC). Recent daily station observations (temperature, pressure, precipitation,sunshine duration, etc.) for Germany, quality control not completed yet, version recent, 2020, accessed 2020-07-12. URL https://opendata.dwd.de/climate_environment/CDC/observations_germany/climate/daily/kl/recent/.

[14] Simone Dohle, Tobias Wingen, and Mike Schreiber. Acceptance and adoption of protective measures during the COVID-19 pandemic: The role of trust in politics and trust in science, May 2020. URL osf.io/w52nv.

[15] Nico Dragano, Christoph J. Rupprecht, Olga Dortmann, Maria Scheider, and Morten Wahren- dorf. Higher risk of COVID-19 hospitalization for unemployed: an analysis of 1,298,416 health insured individuals in Germany. medRxiv, 2020. doi: 10.1101/2020.06.17.20133918. URL https://www.medrxiv.org/content/early/2020/06/19/2020.06.17.20133918.

[16] Maria Effenberger, Andreas Kronbichler, Jae Il Shin, Gert Mayer, Herbert Tilg, and Paul Perco. Association of the COVID-19 pandemic with Internet Search Volumes: A Google Trends(TM) Analysis. International journal of infectious diseases: IJID: official publication of the International Society for Infectious Diseases, 95:192–197, April 2020. ISSN 1878-3511 1201-9712 1201-9712. doi: 10.1016/j.ijid.2020.04.033.

[17] Samuel Engle, John Stromme, and Anson Zhou. Staying at home: mobility effects of COVID-19. Available at SSRN, 2020. URL http://dx.doi.org/10.2139/ssrn.3565703.

[18] James H. Fowler, Seth J. Hill, Nick Obradovich, and Remy Levin. The Effect of Stay-at-Home Orders on COVID-19 Cases and Fatalities in the United States. medRxiv, 2020. doi: 10.1101/2020.04.13.20063628. URL https://www.medrxiv.org/content/early/2020/05/12/2020.04.13.20063628.

[19] Oguzhan Gencoglu and Mathias Gruber. Causal modeling of twitter activity during COVID-19. medRxiv, 2020. doi: 10.1101/2020.05.16.20103903. URL https://www.medrxiv.org/content/early/2020/05/20/2020.05.16.20103903.

[20] Google LLC. Google COVID-19 community mobility reports, 2020, accessed 2020-06-25. URL https://www.google.com/covid19/mobility/.

[21] Google LLC. Google Trends, search term “corona”, 2020, accessed 2020-06-25. URL https://www.google.com/trends.

[22] Sander Greenland, Judea Pearl, and James M. Robins. Causal Diagrams for Epidemiologic Research. Epidemiology, 10(1):37–48, 1999. ISSN 1044-3983. URL https://journals.lww.com/epidem/Fulltext/1999/01000/Causal_Diagrams_for_Epidemiologic_Research.8.aspx.

[23] Sander Greenland, James M. Robins, and Judea Pearl. Confounding and Collapsibility in Causal Inference. Statistical Science, 14(1):29–46, 1999. ISSN 08834237. URL http://www.jstor.org/stable/2676645.

[24] Wei-jie Guan, Zheng-yi Ni, Yu Hu, Wen-hua Liang, Chun-quan Ou, Jian-xing He, Lei Liu, Hong Shan, Chun-liang Lei, David S.C. Hui, Bin Du Lan-juan Li, Guang Zeng, Kwok-Yung Yuen, Ru-chong Chen, Chun-li Tang, Tao Wang, Ping-yan Chen, Jie Xiang, Shi-yue Li, Jin-lin Wang, Zi-jing Liang, Yi-xiang Peng, Li Wei, Yong Liu, Ya-hua Hu, Peng Peng, Jian-ming Wang, Ji-yang Liu, Zhong Chen, Gang Li, Zhi-jian Zheng, Shao-qin Qiu, Jie Luo, Chang-jiang Ye, Shao-yong Zhu, and Nan-shan Zhong. Clinical characteristics of coronavirus disease 2019 in China. New England Journal of Medicine, 2020. doi: 10.1056/NEJMoa2002032.

[25] Leonard Henckel, Emilija Perković and Marloes H. Maathuis. Graphical Criteria for Efficient Total Effect Estimation via Adjustment in Causal Linear Models. arXiv e-prints, art. 1907.02435, July 2019.

[26] Thomas S. Higgins, Arthur W. Wu, Dhruv Sharma, Elisa A. Illing, Kolin Rubel, and Jonathan Y. Ting. Correlations of Online Search Engine Trends With Coronavirus Disease (COVID-19) Incidence: Infodemiology Study. JMIR public health and surveillance, 6(2):e19702, May 2020. ISSN 2369-2960 2369-2960. doi: 10.2196/19702.

[27] Joseph M. Hilbe and William H. Greene. 4 − Count Response Regression Models. In C.R. Rao, J.P. Miller, and D.C. Rao, editors, Essential Statistical Methods for Medical Statistics, pages 104–145. North-Holland, Boston, January 2011. ISBN 978-0-444-53737-9. URL http://www.sciencedirect.com/science/article/pii/B9780444537379500074.

[28] Najaf Iqbal, Zeeshan Fareed, Farrukh Shahzad, Xin He, Umer Shahzad, and Ma Lina. The nexus between COVID-19, temperature and exchange rate in Wuhan city: New findings from partial and multiple wavelet coherence. The Science of the total environment, 729:138916, April 2020. ISSN 1879-1026 0048-9697 0048-9697. doi: 10.1016/j.scitotenv.2020.138916.

[29] Nazrul Islam, Stephen J Sharp, Gerardo Chowell, Sharmin Shabnam, Ichiro Kawachi, Ben Lacey Joseph M Massaro, Ralph B D’Agostino and Martin White. Physical distancing interventions and incidence of coronavirus disease 2019: natural experiment in 149 countries. BMJ, 370, 2020. doi: 10.1136/bmj.m2743. URL https://www.bmj.com/content/370/bmj.m2743.

[30] Mehdi Jahangiri, Milad Jahangiri, and Mohammadamir Najafgholipour. The sensitivity and specificity analyses of ambient temperature and population size on the transmission rate of the novel coronavirus (COVID-19) in different provinces of Iran. The Science of the total environment, 728:138872, April 2020. ISSN 1879-1026 0048-9697 0048-9697. doi: 10.1016/j.scitotenv.2020.138872.

[31] Peter Jüni Martina Rothenbühler Pavlos Bobos Kevin E. Thorpe, Bruno R. da Costa, David N. Fisman, Arthur S. Slutsky, and Dionne Gesink. Impact of climate and public health interventions on the COVID-19 pandemic: A prospective cohort study. Canadian Medical Association Journal, May 2020. ISSN 1488-2329 0820-3946. doi: 10.1503/cmaj.200920.

[32] Markus Kalisch, Martin Mächler Diego Colombo, Marloes H. Maathuis, and Peter Bühlmann. Causal Inference Using Graphical Models with the R Package pcalg. Journal of Statistical Software, 47(11):1–26, 2012. doi: 10.18637/jss.v047.i11. URL http://www.jstatsoft.org/v47/i11/.

[33] William O Kermack and Anderson G McKendrick. Contributions to the mathematical theory of epidemics–i. 1927. Bulletin of mathematical biology, 53(1-2):33—55, 1991. doi: 10.1007/bf02464423.

[34] Moritz U. G. Kraemer, Chia-Hung Yang, Bernardo Gutierrez, Chieh-Hsi Wu, Brennan Klein, David M. Pigott, Louis du Plessis, Nuno R. Faria, Ruoran Li, William P. Hanage, John S. Brownstein, Maylis Layan, Alessandro Vespignani, Huaiyu Tian, Christopher Dye, Oliver G. Pybus, and Samuel V. Scarpino. The effect of human mobility and control measures on the COVID-19 epidemic in China. Science (New York, N.Y.), 368(6490):493–497, May 2020. ISSN 1095-9203 0036-8075 0036-8075. doi: 10.1126/science.abb4218.

[35] Shengjie Lai, Nick W Ruktanonchai, Liangcai Zhou, Olivia Prosper, Wei Luo, Jessica R Floyd, Amy Wesolowski, Mauricio Santillana, Chi Zhang, Xiangjun Du, Hongjie Yu, and Andrew J Tatem. Effect of non-pharmaceutical interventions to contain COVID-19 in China. Nature, May 2020. ISSN 0028-0836. doi: 10.1038/s41586-020-2293-x. URL https://doi.org/10.1038/s41586-020-2293-x.

[36] Arielle Lasry, Daniel Kidder, Marisa Hast, Jason Poovey, Gregory Sunshine, Kathryn Winglee, Nicole Zviedrite, Faruque Ahmed, Kathleen A Ethier, CDC Public Health Law Program, New York City Department of Health and Mental Hygiene, Louisiana Department of Health, Public Health - Seattle & King County, San Francisco COVID-19 Response Team, Alameda County Public Health Department, San Mateo County Health Department, and Marin County Division of Public Health. Timing of community mitigation and changes in reported COVID-19 and community mobility - four U.S. metropolitan areas, February 26-April 1, 2020. MMWR. Morbidity and mortality weekly report, 69(15):451—457, April 2020. ISSN 0149-2195. doi: 10.15585/mmwr.mm6915e2. URL https://doi.org/10.15585/mmwr.mm6915e2.

[37] Cuilian Li, Li Jia Chen, Xueyu Chen, Mingzhi Zhang, Chi Pui Pang, and Haoyu Chen. Retrospective analysis of the possibility of predicting the COVID-19 outbreak from Internet searches and social media data, China, 2020. Euro surveillance: bulletin Europeen sur les maladies transmissibles = European communicable disease bulletin, 25(10), March 2020. ISSN 1560-7917 1025-496X 1025-496X. doi: 10.2807/1560-7917.ES.2020.25.10.2000199.

[38] Yu-Hsuan Lin, Chun-Hao Liu, and Yu-Chuan Chiu. Google searches for the keywords of “wash hands” predict the speed of national spread of COVID-19 outbreak among 21 countries. Brain, behavior, and immunity, April 2020. ISSN 1090-2139 0889-1591 0889-1591. doi: 10.1016/j.bbi.2020.04.020.

[39] Kevin Linka, Mathias Peirlinck, Francisco Sahli Costabal, and Ellen Kuhl. Outbreak dynamics of COVID-19 in Europe and the effect of travel restrictions. Computer methods in biomechanics and biomedical engineering, pages 1–8, May 2020. ISSN 1476-8259 1025-5842. doi: 10.1080/10255842.2020.1759560.

[40] Jiangtao Liu, Ji Zhou, Jinxi Yao, Xiuxia Zhang, Lanyu Li, Xiaocheng Xu, Xiaotao He, Bo Wang, Shihua Fu, Tingting Niu, Jun Yan, Yanjun Shi, Xiaowei Ren, Jingping Niu, Weihao Zhu, Sheng Li, Bin Luo, and Kai Zhang. Impact of meteorological factors on the COVID-19 transmission: A multi-city study in China. The Science of the total environment, 726:138513, April 2020. ISSN 1879-1026 0048-9697 0048-9697. doi: 10.1016/j.scitotenv.2020.138513.

[41] Amaryllis Mavragani. Tracking COVID-19 in Europe: Infodemiology Approach. JMIR public health and surveillance, 6(2):e18941, April 2020. ISSN 2369-2960 2369-2960. doi: 10.2196/18941.

[42] Mattia Mazzoli, David Mateo, Alberto Hernando, Sandro Meloni, and Jose Javier Ramasco. Effects of mobility and multi-seeding on the propagation of the COVID-19 in Spain. medRxiv, page 2020.05.09.20096339, January 2020. doi: 10.1101/2020.05.09.20096339. URL http://medrxiv.org/content/early/2020/05/18/2020.05.09.20096339.abstract.

[43] Timo Mitze, Reinhold Kosfeld, Johannes Rode, and Klaus Wälde. Face masks considerably reduce COVID-19 cases in Germany: A synthetic control method approach. IZA Discussion Papers 13319, Institute of Labor Economics (IZA), 2020. URL https://EconPapers.repec.org/ RePEc:iza:izadps:dp13319.

[44] S. Openshaw. Ecological Fallacies and the Analysis of Areal Census Data. Environment and Planning A: Economy and Space, 16(1):17–31, 1984. doi: 10.1068/a160017. URL https://doi.org/10.1068/a160017.

[45] World Health Organization et al. Transmission of SARS-CoV-2: implications for infection prevention precautions: Scientific Brief, 09 July 2020. Technical report, World Health Organization, 2020.

[46] Judea Pearl. Causality. Cambridge University Press, Cambridge, 2009. ISBN 978-0-521-89560-6. URL https://www.cambridge.org/core/books/causality/B0046844FAE10CBF274D4ACBDAEB5F5B.

[47] Judea Pearl and Elias Bareinboim. External validity: From do-calculus to transportability across populations. Statistical Science, 29(4):579–595, Nov 2014. ISSN 0883-4237. doi: 10.1214/14-sts486. URL http://dx.doi.org/10.1214/14-STS486.

[48] Emilija Perković Johannes Textor, Markus Kalisch, and Marloes H. Maathuis. A Complete Generalized Adjustment Criterion. arXiv e-prints, art. 1507.01524, July 2015.

[49] Emilija PerkovićJohannes Textor, Markus Kalisch, and Marloes H. Maathuis. Complete graphical characterization and construction of adjustment sets in Markov equivalence classes of ancestral graphs. The Journal of Machine Learning Research, 18(1):8132–8193, 2017.

[50] Hongchao Qi, Shuang Xiao, Runye Shi, Michael P. Ward, Yue Chen, Wei Tu, Qing Su, Wenge Wang, Xinyi Wang, and Zhijie Zhang. COVID-19 transmission in Mainland China is associated with temperature and humidity: A time-series analysis. The Science of the total environment, 728:138778, April 2020. ISSN 1879-1026 0048-9697 0048-9697. doi: 10.1016/j.scitotenv.2020.138778.

[51] R Core Team. R: A Language and Environment for Statistical Computing. R Foundation for Statistical Computing, Vienna, Austria, 2019. URL https://www.R-project.org/.

[52] Robert Koch-Institut (RKI). Fallzahlen in Deutschland (COVID-19), 2020, accessed 2020-07-12. URL https://www.rki.de/DE/Content/InfAZ/N/Neuartiges_Coronavirus/Fallzahlen.html.

[53] S. Schipf, S. Knüppel, J. Hardt, and A. Stang. Directed Acyclic Graphs (DAGs) – Die Anwendung kausaler Graphen in der Epidemiologie. Gesundheitswesen, 73(12):888–892, December 2011. ISSN 0941-3790. doi: 10.1055/s-0031-1291192.888.

[54] Peng Shi, Yinqiao Dong, Huanchang Yan, Chenkai Zhao, Xiaoyang Li, Wei Liu, Miao He, Shixing Tang, and Shuhua Xi. Impact of temperature on the dynamics of the COVID-19 outbreak in China. The Science of the total environment, 728:138890, April 2020. ISSN 1879-1026 0048-9697 0048-9697. doi: 10.1016/j.scitotenv.2020.138890.

[55] Marcos Felipe Falcão Sobral Gisleia Benini Duarte, Ana Iza Gomes da Penha Sobral, Marcelo Luiz Monteiro Marinho, and André de Souza Melo. Association between climate variables and global transmission of SARS-CoV-2. Science of the Total Environment, 729:138997, April 2020. ISSN 1879-1026 0048-9697 0048-9697. doi: 10.1016/j.scitotenv.2020.138997.

[56] Peter Spirtes, Clark N Glymour, Richard Scheines, and David Heckerman. Causation, prediction, and search. MIT press, 2000. ISBN 0-262-19440-6.

[57] Johannes Textor, Benito van der Zander, Mark S Gilthorpe, Maciej Liśkiewicz and George TH Ellison. Robust causal inference using directed acyclic graphs: the R package ‘dagitty’. International Journal of Epidemiology, 45(6):1887–1894, January 2017. ISSN 0300-5771. doi: 10.1093/ije/dyw341. URL https://doi.org/10.1093/ije/dyw341.

[58] Ramadhan Tosepu, Joko Gunawan, Devi Savitri Effendy, La Ode Ali Imran Ahmad, Hariati Lestari, Hartati Bahar, and Pitrah Asfian. Correlation between weather and Covid-19 pandemic in Jakarta, Indonesia. The Science of the total environment, 725:138436, April 2020. ISSN 1879-1026 0048-9697. doi: 10.1016/j.scitotenv.2020.138436.

[59] W. N. Venables and B. D. Ripley. Modern Applied Statistics with S. Springer, New York, fourth edition, 2002. URL http://www.stats.ox.ac.uk/pub/MASS4. ISBN 0-387-95457-0.

[60] Abigail Walker, Claire Hopkins, and Pavol Surda. Use of Google Trends to investigate loss-of-smell–related searches during the COVID-19 outbreak. International Forum of Allergy & Rhinology, 10(7):839–847, 2020. doi: 10.1002/alr.22580. URL https://onlinelibrary.wiley.com/doi/abs/10.1002/alr.22580.

[61] Mao Wang, Aili Jiang, Lijuan Gong, Lina Luo, Wenbin Guo, Chuyi Li, Jing Zheng, Chaoyong Li, Bixing Yang, Jietong Zeng, Youping Chen, Ke Zheng, and Hongyan Li. Temperature significant change COVID-19 transmission in 429 cities. medRxiv, 2020. doi: 10.1101/2020.02.22.20025791. URL https://www.medrxiv.org/content/early/2020/02/25/2020.02.22.20025791.

[62] WHO Team. Report of the WHO-China joint mission on coronavirus disease 2019 (COVID-19), 2020, accessed 2020-06-25. URL https://www.who.int/publications-detail/report-of-the-who-china-joint-mission-on-coronavirus-disease-2019-(covid-19).

[63] Yu Wu, Wenzhan Jing, Jue Liu, Qiuyue Ma, Jie Yuan, Yaping Wang, Min Du, and Min Liu. Effects of temperature and humidity on the daily new cases and new deaths of COVID-19 in 166 countries. Science of The Total Environment, 729:139051, 2020. ISSN 0048-9697. doi: https://doi.org/10.1016/j.scitotenv.2020.139051. URL http://www.sciencedirect.com/science/article/pii/S0048969720325687.

[64] Jingui Xie and Yongjian Zhu. Association between ambient temperature and COVID-19 infection in 122 cities from China. The Science of the total environment, 724:138201, July 2020. ISSN 1879-1026 0048-9697 0048-9697. doi: 10.1016/j.scitotenv.2020.138201.

[65] Chenfeng Xiong, Songhua Hu, Mofeng Yang, Hannah N Younes, Weiyu Luo, Sepehr Ghader, and Lei Zhang. Data-Driven Modeling Reveals the Impact of Stay-at-Home Orders on Human Mobility during the COVID-19 Pandemic in the U.S. arXiv e-prints, art. 2005.00667, May 2020.

[66] Ye Yao, Jinhua Pan, Zhixi Liu, Xia Meng, Weidong Wang, Haidong Kan, and Weibing Wang. No association of COVID-19 transmission with temperature or UV radiation in Chinese cities. The European respiratory journal, 55(5), May 2020. ISSN 1399-3003 0903-1936 0903-1936. doi: 10.1183/13993003.00517-2020.

[67] Xiaoling Yuan, Jie Xu, Sabiha Hussain, He Wang, Nan Gao, and Lanjing Zhang. Trends and Prediction in Daily New Cases and Deaths of COVID-19 in the United States: An Internet Search-Interest Based Model. Exploratory research and hypothesis in medicine, 5(2):1–6, April 2020. ISSN 2472-0712 2472-0712 2472-0712. doi: 10.14218/ERHM.2020.00023.

[68] Wei Ke Zhou, Ai Li Wang, Fan Xia, Yan Ni Xiao, and San Yi Tang. Effects of media reporting on mitigating spread of COVID-19 in the early phase of the outbreak. Mathematical biosciences and engineering: MBE, 17(3):2693–2707, March 2020. ISSN 1551-0018 1547-1063. doi: 10.3934/mbe.2020147.

